# Is physical rehabilitation superior to no physical rehabilitation following total knee arthroplasty? A systematic review and meta-analysis

**DOI:** 10.1101/2020.04.29.20084392

**Authors:** T. Mark-Christensen, C. Juhl, K. Thorborg, T. Bandholm

**Affiliations:** Centre of Health, Department of Rehabilitation, Bornholms Regionskommune, Curdtslund 2,3700 Rønne, Denmark; Department of Physiotherapy and Occupational Therapy, University Hospital of Copenhagen, Herlev and Gentofte, Denmark; Department of Sports Science and Clinical Biomechanics, University of Southern Denmark, Odense, Denmark; Physical Medicine and Rehabilitation Research-Copenhagen (PMR-C); Department of Physical and Occupational Therapy; Clinical Research Centre; Department of Orthopaedic Surgery, Hvidovre Hospital, University of Copenhagen, Hvidovre Denmark; Sports Orthopaedic Research Centre – Copenhagen (SORC-C), Arthroscopic Centre Amager, Department of Orthopaedic Surgery, Copenhagen University Hospital, Copenhagen, Amager-Hvidovre, Denmark

## Abstract

**Introduction:** Physical rehabilitation is widely used following total knee arthroplasty (TKA), while no consensus has been reached regarding the optimal dosage, frequency and modality. Before a standardized protocol can be established, it is important to investigate the true efficacy for physical rehabilitation following TKA.

**Objective:** To examine if physical rehabilitation is superior to no physical rehabilitation following total knee arthroplasty in terms of patient-reported outcomes for function and pain.

**Methods:** The search strategy was conducted in five databases (MEDLINE, Embase, Cinahl, CENTRAL and SPORTDiscus) with eligibility criteria for inclusion being: Randomised controlled trials comparing patients receiving physical rehabilitation with a group receiving *no* physical rehabilitation following unilateral TKA. Potential sources of bias were assessed according to the Cochrane Risk of Bias Tool. The Consensus on Exercise Reporting Template (CERT) was used to extract and report exercise interventions from the included trials.

**Results:** Only two of 3254 identified trials fulfilled the eligibility criteria and were included. The two trials were judged as being of moderate to good methodological quality, but lacking in the reporting of their exercise interventions. Difference in outcome measurements and interventions makes between-study comparison inconclusive.

**Conclusions:** No conclusion regarding the superiority of physical rehabilitation to no-intervention following TKA can be drawn from the results of this systematic review. Further research is required to establish the true effectiveness of physical rehabilitation following TKA.

Systematic review registration number: PROSPERO 2018 CRD42018094785.

## Introduction

Physical rehabilitation following total knee arthroplasty (TKA) is widely used after discharge from hospital, as it seems to speed up the functional recovery following surgery ^1^. However, physical rehabilitation components after discharge still vary significantly in content, duration and intensity, where current TKA-rehabilitation practices are ranging from a single session of professional advice while still in hospital to several weeks of intensive in- or outpatient physical rehabilitation to facilitate a full recovery ^2–4^. A survey undertaken in 2012 in England and Wales found 16 of the 23 included orthopaedic centres referred patients to physical rehabilitation as a standard after TKA, while six centres did not offer routine physical rehabilitation after discharge ^5^. In Denmark, the referral percentage to post-discharge physical rehabilitation following TKA varies from 15% to 100%, depending on geographic region ^6^ and patients with no formal post-discharge physical rehabilitation receive only inpatient rehabilitation with instructions of post-discharge home-exercises. Several systematic reviews of randomized controlled trials have concluded that no single setting (clinic- or home-based, water- or land based, supervised or unsupervised) appears to be superior to other interventions across a range of outcome measures following TKA ^1,7,8^. As no single intervention has proven superiority to other interventions (e.g. all interventions seem equally effective), is has become necessary to investigate the true efficacy of physical rehabilitation following TKA. This can be done by comparing physical rehabilitation superiority to no-intervention.

Artz et al. (2015) analysed seven trials comparing physical rehabilitation to “no or minimal intervention” in terms of physical function (five trials), pain (four trials) and range of motion (five trials) ^1^. However, it is important to distinguish between minimal and no-intervention, as minimal intervention suggests that *some* effort has been made to rehabilitate patients following TKA (for example by providing the patient with an exercise booklet). No-intervention does, however, not include any intervention designed to restore function or reduce pain in the knee. Only by comparing *some* physical rehabilitation to *no* physical rehabilitation, the true efficacy of physical rehabilitation following TKA can be determined. *Some* physical rehabilitation is defined as: physical activity and exercise designed and prescribed for restoring normal function or reducing pain caused by disease or injury. *No* physical rehabilitation means none of the above, but could include encouragement to stay active and continue life as usual.

Based on the above, the research question of this systematic review and meta-analysis is: Is physical rehabilitation superior to no physical rehabilitation following total knee arthroplasty in terms of the patient-reported outcomes for function and pain? The research question that fostered the objective was developed using the PICOT approach ^9^ and FINER criteria ^10^ to ensure scientific and clinically relevance. Specifically, input regarding relevance of the research question was collected from the following stakeholder; patients, orthopaedic surgeons, physiotherapists and policy-makers. In summary, these stakeholders commented that they were very interested in finding out whether or not postoperative physical rehabilitation is superior to no physical rehabilitation following total knee arthroplasty.

### Objective

The primary objective of this systematic review and meta-analysis was to examine if physical rehabilitation is superior to no physical rehabilitation following total knee arthroplasty in terms of patient-reported outcomes for function and pain.

## Methods

The systematic review and meta-analysis was performed following the guidelines from the Cochrane Library ^11^ and reported according to the Preferred Reporting Items for Systematic Reviews and Meta-analyses (PRISMA) recommendations ^12^. This study protocol was registered in the PROSPERO database (international prospective register of systematic reviews) before commencement, based on the protocol manuscript following the PRISMA-P guidelines ^13^. Trial registration: CRD42018094785 (URL: https://www.crd.york.ac.uk/prospero/display_record.php?ID=CRD42018094785)

### Eligibility criteria

<u>Inclusion criteria:</u>

1. Randomised controlled trials comparing patients receiving physical rehabilitation with a group receiving no physical rehabilitation following a unilateral total knee arthroplasty.
2. Interventions commencing at a pre-specified time after discharge (no longer than three months after index surgery to reflect clinical practice).
3. Included trials had included at least one of the following outcomes: continuous measure of pain, patient-reported measure of physical function, performance-based measure of function, or a patient global assessment measurement, as recommended by Osteoarthritis Research Society International (OMERACT-OARSI) ^14,15^.
4. The primary intervention had to be physical rehabilitation. Other interventions (such as electrical stimulation, acupuncture, continuous passive motion) were only allowed as adjunct to physical rehabilitation (meaning less than 20% of the rehabilitation).

Trials that did not fulfil the above-mentioned criteria were excluded.

### Search strategy

PROSPERO was screened for any on-going systematic reviews of relevance for this review. The search strategy was conducted in five databases (MEDLINE, Embase, Cinahl, CENTRAL and SPORTDiscus). Each database was searched using a search strategy with keywords and text words (see appendix for the detailed search strategy), combining the search focus 1) total knee arthroplasty or, - replacement, and 2) physical exercise, and rehabilitation.

Within a search focus the search words were combined with OR and the results between the search focus were combined with AND. Finally, the search was combined with Cochrane Highly Sensitive Search Strategy for identifying randomized trials. Previous published systematic reviews published within the last 5 years were manually checked for relevant studies, not identified in the main search strategy.

### Study selection

The result from the main search strategy was downloaded to reference-managing software, where duplicates were removed. Two authors (TMC and TB) independently selected eligible studies going through title and abstract. Studies judged eligible by at least one review author were evaluated by the same two review authors (TMC and TB) in full text based on the inclusion criteria (first study selection step).

Trial data (authors, year of publication, study design, number of participants, outcome measures, trial duration and trial registration), patient-related data (gender, age, BMI), and outcome data (baseline and follow up measures of patient-reported function and pain, performance-based measures of function and global patient outcome assessment) was extracted by two review authors (TMC and TB) independently from the included trials. The Consensus on Exercise Reporting Template (CERT) was used to extract and report exercise interventions from the included trials ^16^. Consensus was reached by discussion. If relevant data were missing or unclear within an included trial, the corresponding author of the trial was contacted for further clarification.

### Outcome

The primary outcome was change in patient-reported outcomes for function and pain from baseline to first outcome assessment after the intervention (≤6 months, primary analysis). Patient-reported function can be measured through e.g. Knee injury and Osteoarthritis Outcome Score (KOOS), Western Ontario and McMaster Universities Osteoarthritis Index (WOMAC), Knee Society Clinical Rating System (KSS), Oxford Knee Score (OKS) or by a global assessment of function. Patient global assessment of function can be measured by a standard question by a single Likert or visual analogue scale (VAS) metric, asking a standard question, for example “How would you rate your current level of function during your usual activities of daily living?” ^14^. Pain can be measured by VAS, numeric analogue scale (NRS), Likert scale of severity, or through subscales in patient-questionnaires as KOOS, WOMAC and KSS ^17^.

Secondary outcome measurements were changes in performance-based measure of function from baseline to outcome assessments. Performance-based measure of function include objective assessments as recommended by OMERACT-OARSI ^14,15^, e.g. 30-seconds Sit to Stand, 4 × 10 m fast-paced walk test.

### Risk of bias

Potential sources of bias was assessed according to the Cochrane Risk of Bias Tool version 2 (RoB 2) ^18^ and presented as a “Risk of bias summary” including five domains; (1) bias arising from the randomization process, (2) bias due to deviations from intended interventions, (3) bias due to missing outcome data, (4) bias in measurement of the outcome, and (5) bias in selection of the reported result. Each of the mentioned domains was evaluated as to whether they were considered adequate (low risk of bias), some concern (some concern of risk of bias) or inadequate (high risk of bias) ^18^.

Two review authors (TMC and TB) independently evaluated the risk of bias domains and disagreement between the reviewers was solved either by consensus or involving a third reviewer (KT).

### Data synthesis

Only a small number of studies could be included in this study. An overview of the results was therefore firstly presented in a narrative way for each of the individual studies and secondly, as an overview of the results within each of the relevant outcome domains.

## Results

The review progress is summarised as a flow diagram (figure 1). Out of 3254 identified studies, two randomised controlled trials were found eligible for inclusion.

**Figure 1.**
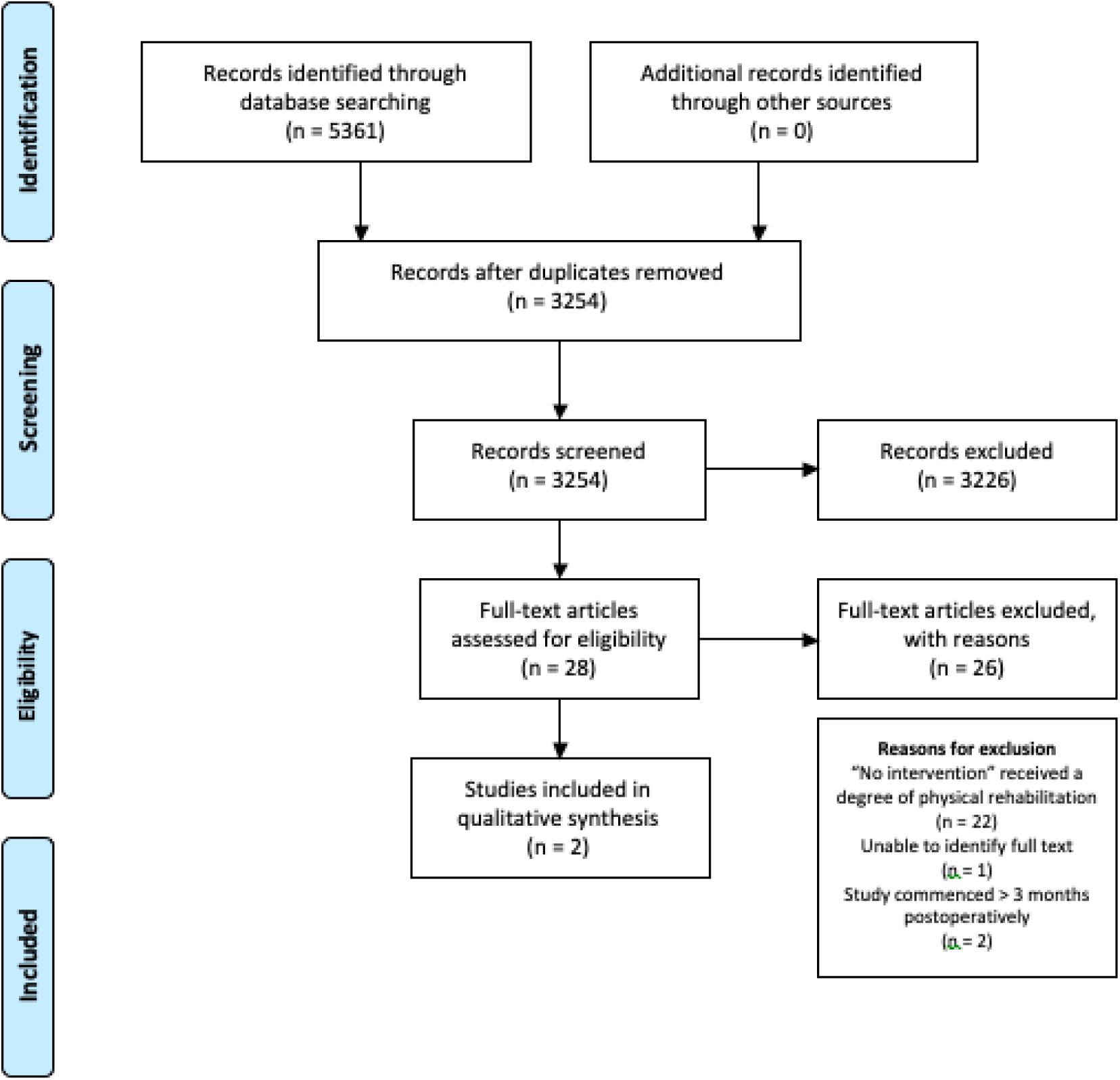
Flow diagram of review progress and study selection.

### Summery of included trials

The trial by Evgeniadis et al. (2008) ^19^ investigated the effect of a rehabilitation program before and after TKA using a 3-arm randomized controlled trial design, dividing the patients into a only preoperative exercise group (3 week program), a only postoperative exercise group (8 week program) or a control group, who had no treatment.

Monticone et al. (2013) ^20^ investigated the effectiveness of a 6 month home-based functional and cognitive-behavioural exercise program compared to acontrol group, who were given general advise to stay active after discharge from a rehabilitation unit after TKA.

Trial characteristics for both trials are presented in table 1.

**Table 1.**
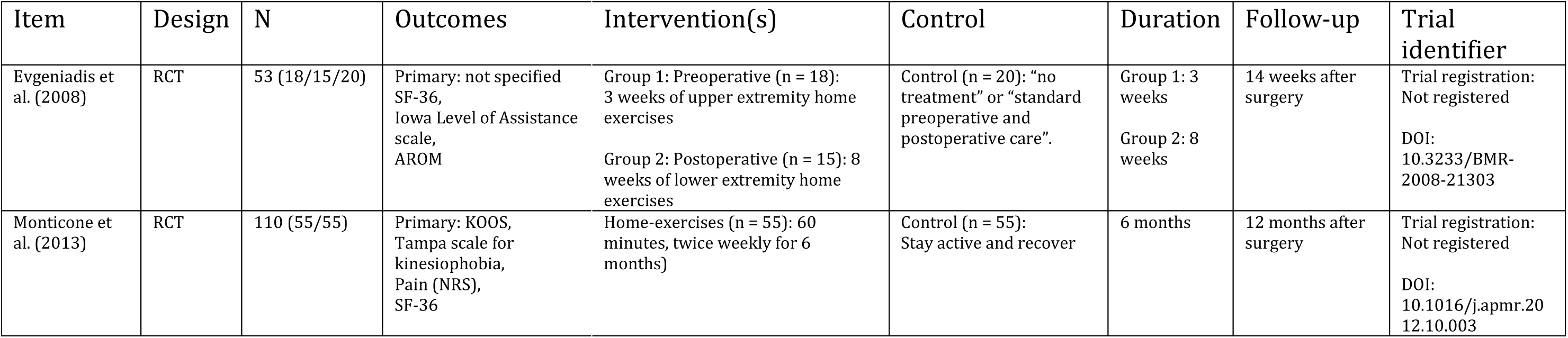
Trial characteristics of included trials

### Risk of bias

Results from the methodological quality assessment using the Cochrane Risk of Bias Tool version 2 (RoB 2) ^18^ are presented in table 2. While Monticone et al. (2013) were assessed to have low risk of bias, the trial by Evgeniadis et al. (2008) presented some concerns of risk of bias, including small sample sizes, noteworthy and uneven number of dropouts and risk of selective reporting. Each trials exercise intervention is described in detail using “The Consensus on Exercise Reporting Template” (CERT) ^16^ (see appendix). The reporting of the exercise interventions in the two included trials is generally poor and lack important intervention-details, making an intervention replication and comparison difficult.

**Table 2.**
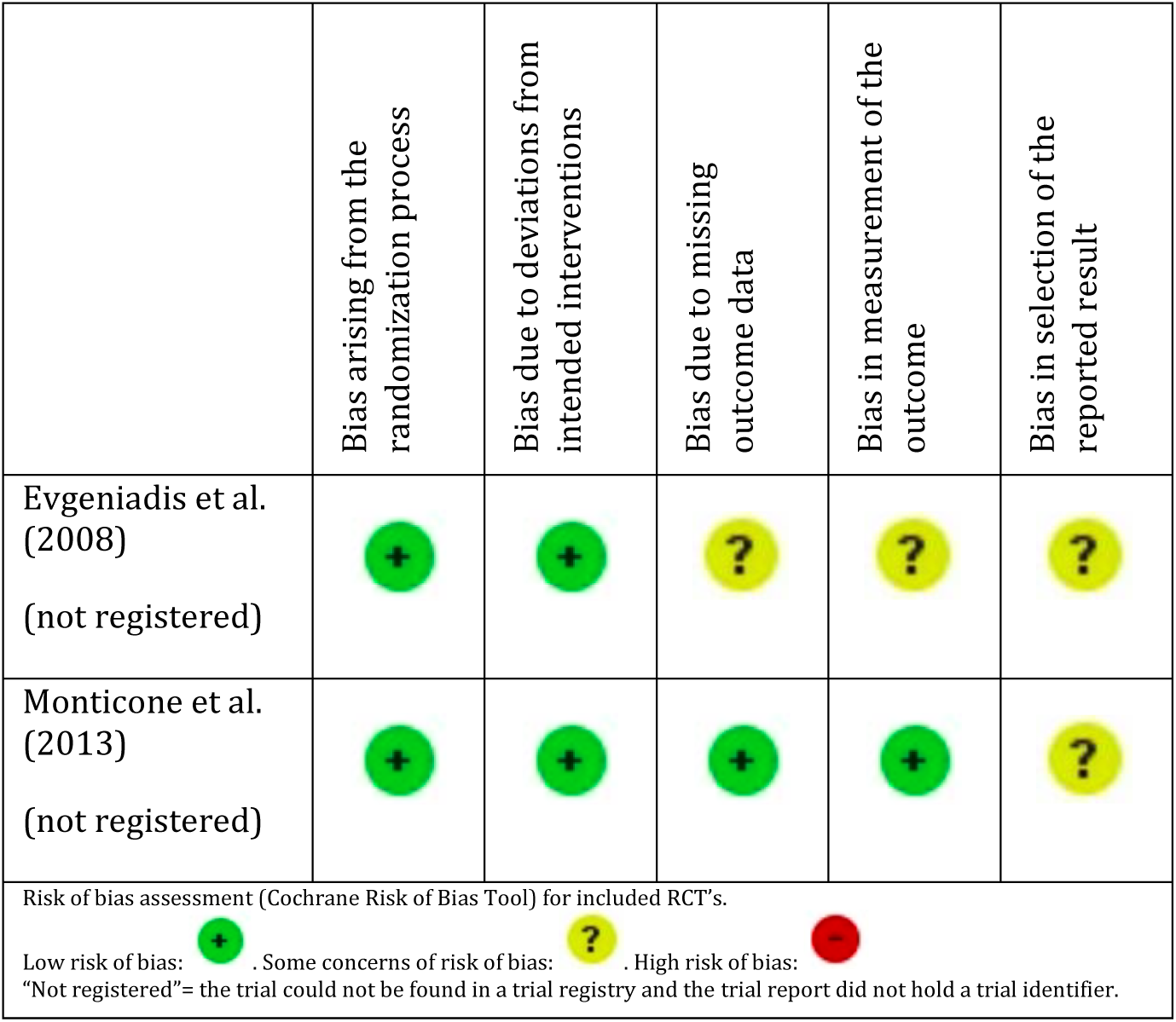
Risk of bias summary.

### Results from the individual studies

As the objective of this systematic review was to examine if physical rehabilitation is superior to no physical rehabilitation *following* TKA, the data from the preoperative intervention group in the trial by Evgeniadis et al. (2008) was not included in this review.

### Intervention focus

The trial by Evgeniadis et al. (2008) compared a physical therapist and orthopaedist-supervised, eight-week home exercise program to a control group, who followed the standard preoperative and postoperative care, which included an inpatient rehabilitation program. The control group did not receive any pre- or postoperative physical rehabilitation compared to the other two intervention groups. After hospital discharge, the intervention group received a therapeutic exercise program with emphasis on strengthening their lower extremities. The program consisted of exercise from supine, lateral and prone position, sitting and straight posture with the aim to strengthen anterior and posterior tibial muscles, anterior and posterior femoral muscles and the femoral abductors.

Monticone et al. (2013) compared an intervention group, who continued the functional exercise learned during hospitalization for 6 months, to a control group, who were advised to stay active and gradually recover to their usual activities. The intervention group performed functional exercises 60 minutes twice-weekly, including walking training, functional task-oriented exercises (moving from sitting to standing, ascending/descending stairs, climbing obstacles), weight-bearing exercises (walking in place, bilateral and unilateral knee flexions while standing) and other exercises aimed at recovering abilities (turning, sudden starts and stops). Sessions of stationary cycling were added to optimize the strength and mobility of the knee.

### Outcome measures

#### Patient reported outcome measures

Evgeniadis et al. (2008) did not report their primary outcome measure. The authors used the SF-36 to assess the effect of the preoperative exercise intervention, while the postoperative exercise intervention was assessed using the Iowa Level of Assistance Scale (ILAS). Differences in active range of motion (AROM) of the hip, knee and tibiotalar joint within the three groups were also investigated.

Monticone et al. (2013) used patient-reported disability as the primary outcome measure. Disability was measured with the Knee Osteoarthritis Outcome Score (KOOS). Also, Tampa Scale for Kinesiophobia (TSK), Numerical Rating Scale (NRS) and the 36-Item Short-Form Health Survey (SF-36) were used as outcome measures. By the end of the 6-month intervention, a mean difference of 14.22 (95%CI: 8.35 to 20.08) in the KOOS-subscale: ADL was found between the exercise and control group, favouring the exercise group. The difference between the groups were still present (mean difference: 11.84 (95%CI: 6.79 to 16.89)) at the 12 months follow up. The remaining subscales revealed a similar between-group difference (mean differences: 10.81 to 13.31). Fear-avoidance behaviour was assessed using the TSK, which also showed results favouring the exercise group (difference in treatment effect between the two groups were found to be 25%). Pain and quality of life also revealed a between-group difference at both follow-ups, favouring the exercise group.

#### Performance-based outcome measures

Evgeniadis et al. (2003) used the ILAS to assess the patients’ functional ability postoperatively. The groups did not appear to difference from the day before surgery until ten weeks after the surgery. By the tenth postoperative week, the exercise group performed favourably in terms of ambulatory independence compared to the control group (mean score 2.79 (SD: 0.64) vs. 4.87 (SD: 0.73)), but at the final follow up four weeks later, there were no between-group difference (0.14 (SD: 0.39) vs. 0.38 (SD: 0.56)).

Monticone et al. (2013) did not use any performance-based outcome measures.

Neither of the two included studies used patient global assessment measurements, nor did they report harms or unintended effects in relation to the allocated groups.

### Synthesis of results

The primary objective of this systematic review was to examine if physical rehabilitation is superior to no physical rehabilitation following TKA in terms of patient-reported outcomes for function and pain. No conclusion can be drawn at this point because only two studies were included. In the trial of Monticone et al. (2013) functional home-exercises (including cognitive-behavioural therapy) appears to be superior to a no-intervention, in terms of the primary outcome, patient-reported disability, with a mean adjusted difference of 14.22 (95%CI: 8.35, 20.08) after the intervention period. The difference is considered to be above the minimal clinical important difference, which is reported to be around 10 points ^21,22^. Secondary outcome measures used in the trial, such as kinesiophobia, pain and quality of life, were also found to be superior in the intervention group compared to no-intervention.

Evgeniadis et al. (2008) found no difference in terms of performance status (measured by SF-36), but a tendency towards a higher score in “mental health” and “health change” in the postoperative exercise group compared to the control group. Ten weeks after surgery, the mean difference in functional ability measured by ILAS, were higher in the postoperative exercise group compared to the preoperative exercise group and the control group (total score of 2.79, compared to 4.65 and 4.78, respectively), but this difference was no longer statistically significant 14 weeks after surgery. AROM were also reported to be significantly better in the postoperative exercise group compared to the other two groups.

## Discussion

To date, there is not enough evidence to either confirm or refute, that physical rehabilitation is superior to no physical rehabilitation following total knee arthroplasty in terms of patient-reported outcomes for function and pain.

While most research on the subject has involved the identification of a superior rehabilitation protocol compared to another or “standard care”, little effort has been made to find the true effectiveness of physical rehabilitation following TKA. A recent systematic review and meta-analysis on participants after TKA compared physiotherapy exercise to “no intervention” ^1^, and found short-term benefits for physical function (SMD −0.37 (95%CI: −0.62 to −0.12: three studies), and pain (SMD −0.45 (95%CI: −0.85 to −0.06: two studies) favouring physical rehabilitation. However, two of the three studies concerning physical function and one of the two concerning pain, included a control group, whom performed some kind of home exercise. While definitely being “minimal intervention”, it is not the same as “no intervention”. Additionally, when analysing the effects in studies with low risk of bias in the previous mentioned review, only one or two trials were included in the analysis. The two studies investigating a true “no intervention” included in the review by Artz et al. (2015) ^1^ are also the two trials included in this systematic review.

A true “no intervention” is on the other hand controversial. While TKA is followed by the declining of muscle strength and functional ability ^23–25^ is seems reasonable to support the recovery from TKA by offering physical rehabilitation. The effectiveness of different modalities has been investigated, and there is compiling evidence that the degree of supervision during physical rehabilitation is not a cofounder in terms of recovery ^1,26^. Prescribing unsupervised home exercises does however usually require some degree of supervised exercise instructions and perhaps a follow-up visit or two within the first couple of months of surgery. Due to its low cost of administration, and equal effectiveness compared to other rehabilitation modalities, unsupervised home exercises seems to be the preferred choice of treatment.

Do patients, who are prescribed an unsupervised home exercise program following TKA, actually exercise at home? Campbell and colleagues ^27^ found that non-compliance is a common problem (and increases over time) in a cohort of patients with knee osteoarthritis, when asked to perform home exercises. The authors raise the question as to whether we should judge effectiveness according to whether an intervention works when compliance is optimal or taking into account variable levels of compliance ^27^. Home exercises following TKA may have an inherent design flaw since adherence is rarely measured and the true effectiveness of the home exercises can therefore be questioned.

There is a need for large, randomized controlled trials investigating the effectiveness of physical rehabilitation following TKA when compared to a true “no intervention” to either establish or disprove the effectiveness of physical rehabilitation following TKA.

### Limitations and strengths

An obvious limitation to this systematic review is the limited number of randomized controlled trials available on the topic. The considerable clinical heterogeneity of the exercise interventions makes it impossible to conclude on the effectiveness of physical rehabilitation compared to a true no-intervention. Furthermore, the risk of bias in one of the two included trials should also be taken into consideration when interpreting the overall result of this review. Despite these limitations, this systematic review has strength in its adherence to the PRISMA guidelines, the PICOT approach and using the FINER criteria. Furthermore, by publishing the protocol for this review, we strengthened the transparency of the research and minimise the risk of bias, namely selective outcome reporting.

This systematic review was first published as preprint in order to determine the order of priority and to increase the transparency of the scientific work ^28^. Sharing this work as preprint before final publication will allow for early evaluation by the scientific community and further improve the quality of the review. This review will be frequently updated to keep the review’s results accurate and up to date. This form of “living systematic review” will enable living, accurate recommendations and create an environment for more dynamic, interlinked synthesis and use of the data ^29^.

## Conclusion

The considerable clinical heterogeneity of the included studies design, exercise interventions and outcomes measurements, makes it difficult to confirm or refute physical rehabilitations superiority over no-intervention following TKA. This systematic review highlights the need for adequately powered RCT’s investigating the effectiveness of physical rehabilitation compared to a true nointervention.

## Data Availability

The authors confirm that the data supporting the findings of this study are available within the article [and/or] its supplementary materials.

## Appendix

### MEDLINE search strategy

1. Arthroplasty, Replacement, Knee [MeSH]
2. Knee Prosthesis [MeSH]
3. Joint Prosthesis [MeSH]
4. Knee [MeSH] OR knee [Title/Abstract]
5. #3 AND #4
6. arthroplast*[Title/Abstract] OR replac* [Title/Abstract] OR prosthe* [Title/Abstract]
7. #4 AND #6
8. #1 OR #2 OR #5 OR #7
9. Exercise [MeSH]
10. Exercise Therapy [MeSH]
11. Rehabilitation [MeSH]
12. Physical Therapy Modalities [MeSH]
13. Resistance Training [MeSH]
14. Cardiorespiratory Fitness [MeSH]
15. Muscle Strength [MeSH]
16. Weight Lifting [MeSH]
17. Sports [MeSH]
18. Physical Endurance [MeSH]
19. Bicycling [MeSH]
20. Walking [MeSH]
21. Exercise* [Title/Abstract]
22. Weight [Title/Abstract] AND Lifting [Title/Abstract]
23. isometric* [Title/Abstract] OR isotonic* [Title/Abstract] OR isokinetic* [Title/Abstract]
24. strength [Title/Abstract] AND #23
25. Treadmill* [Title/Abstract]
26. Bicycle* [Title/Abstract]
27. Cycling [Title/Abstract]
28. Walk* [Title/Abstract]
29. Sport* [Title/Abstract]
30. Endur* [Title/Abstract]
31. #9 OR #10 OR #11 OR #12 OR #13 OR #14 OR #15 OR #16 OR #17 OR #18 OR #19 OR #20 OR #21 OR #22 OR #24 OR #25 OR #26 OR #27 OR #28 OR #29 OR #30
32. Randomized Controlled Trial [Publication Type]
33. Controlled clinical trial [Publication Type]
34. Randomized [Title/Abstract]
35. Randomly [Title/Abstract]
36. Trial [Title/Abstract]
37. Groups [Title/Abstract]
38. #32 OR #33 OR #34 OR #35 OR #36 OR #37
39. **#8 AND #31 AND #38**

**Table XX.**
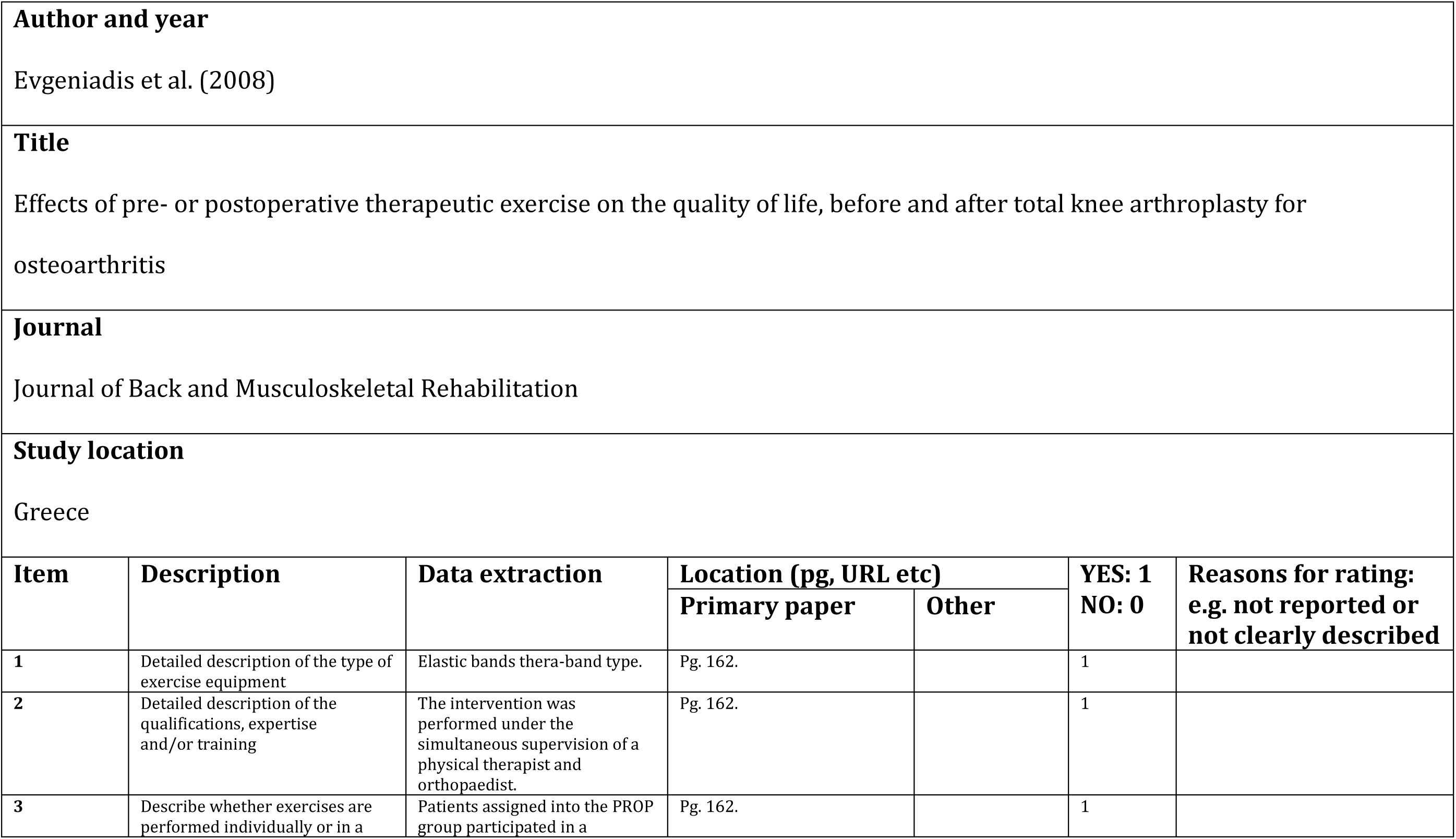
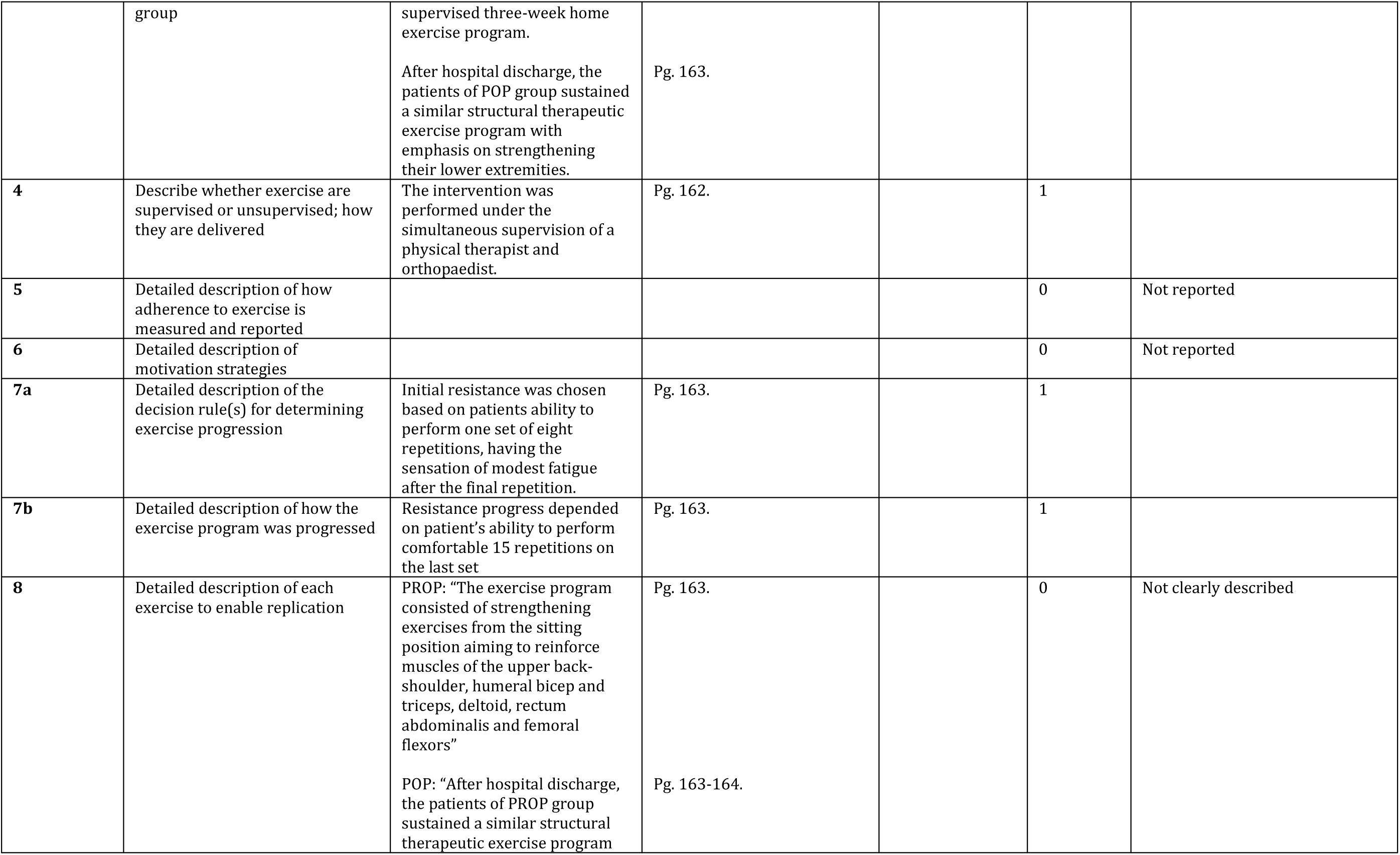
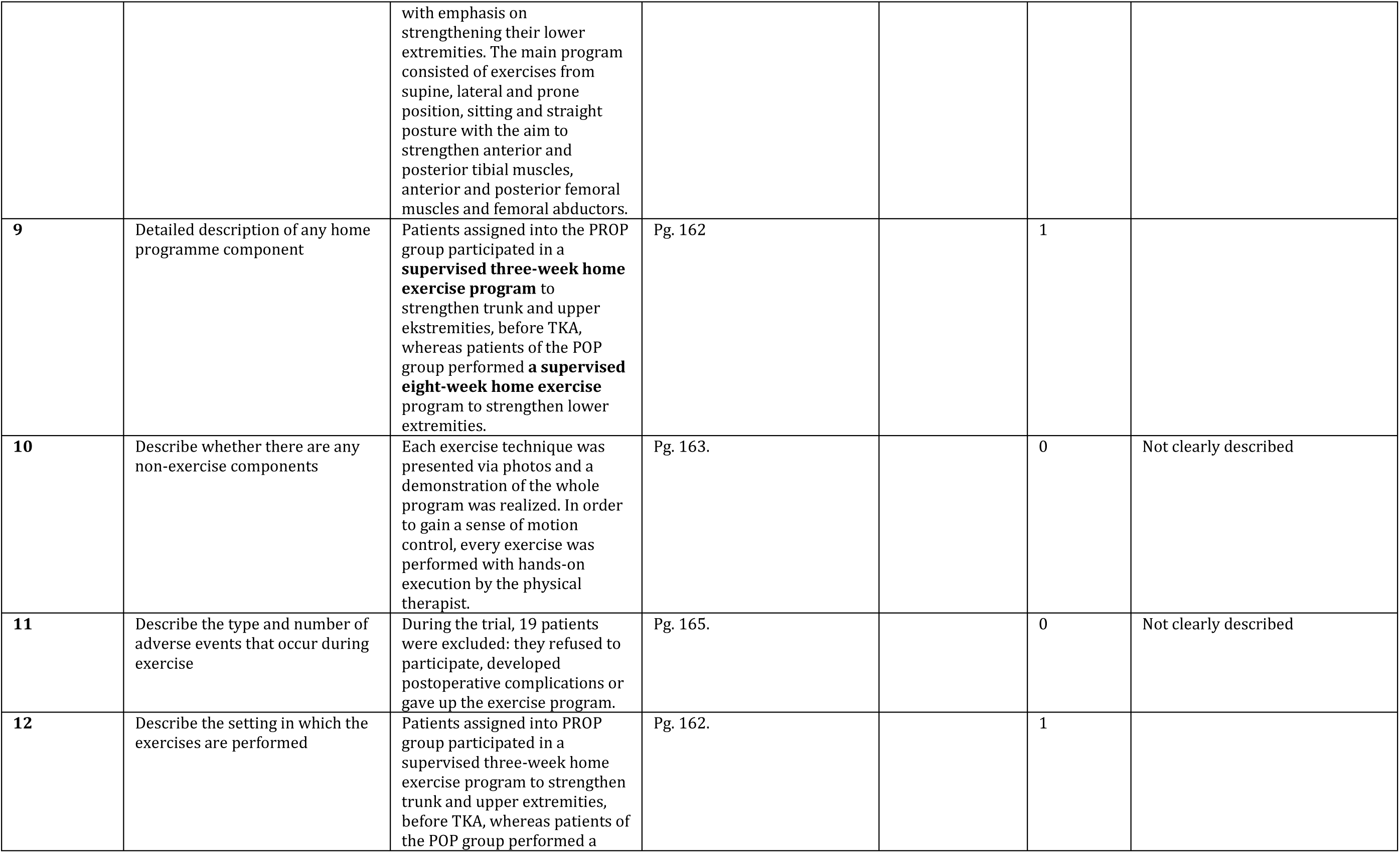
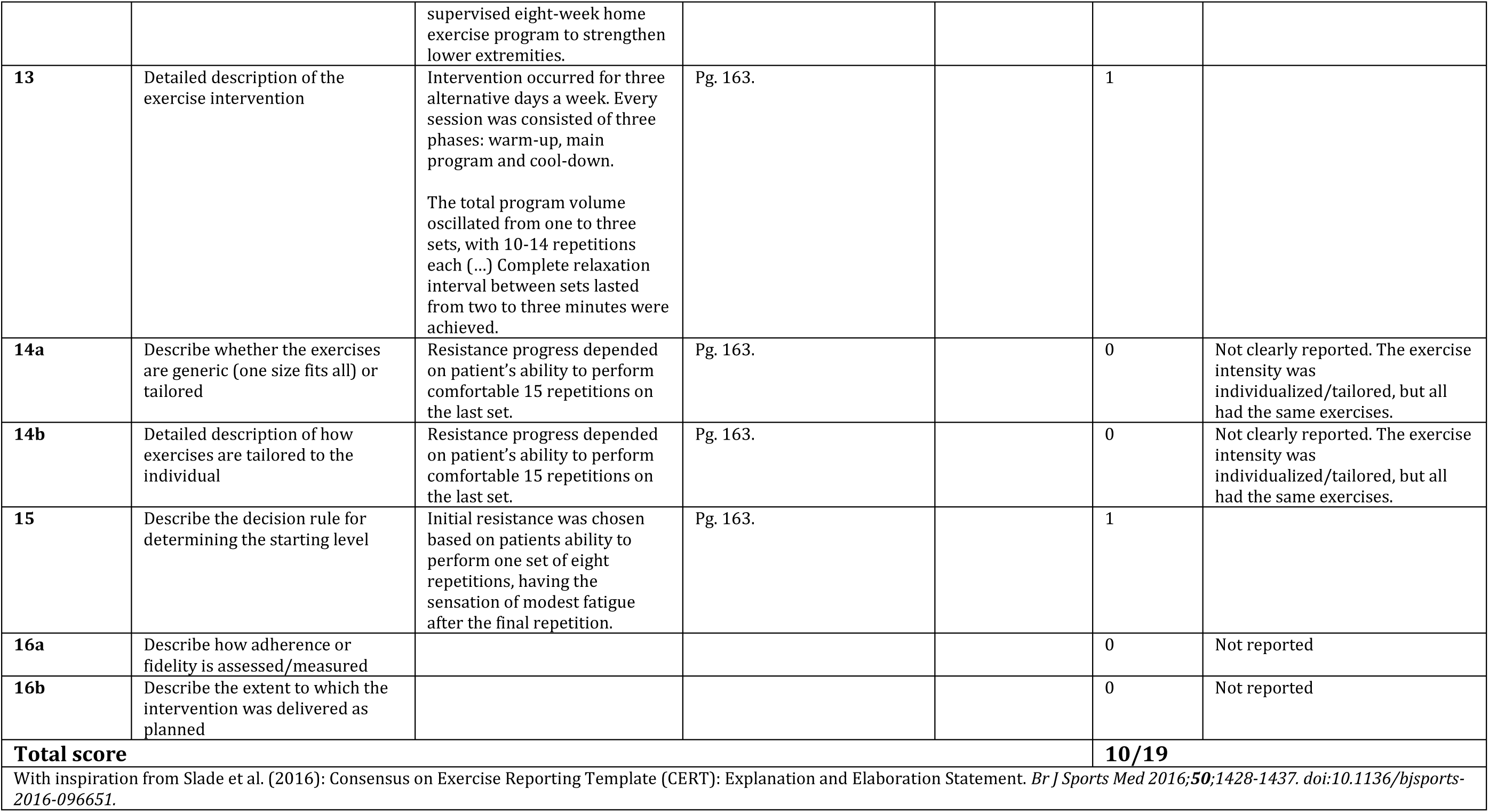
CERT assessment form

**Table XX.** CERT assessment form

| **Author and year** |  |  |  |  |  |  |
| --- | --- | --- | --- | --- | --- | --- |
| Monticone et al. (2013) |  |  |  |  |  |  |
| **Title** |  |  |  |  |  |  |
| Home-based functional exercises aimed at managing kinesiophobia contribute to improving disability and quality of life of patients undergoing total knee arthroplasty: a randomized controlled trial |  |  |  |  |  |  |
| **Journal** |  |  |  |  |  |  |
| Archives of physical medicine and rehabilitation |  |  |  |  |  |  |
| **Study location** |  |  |  |  |  |  |
| Italy |  |  |  |  |  |  |
| Item | Description | Data extraction | Location (pg, URL etc) |  | YES: 1<br>NO: 0 | Reasons for rating:<br>eg 'not reported or<br>not clearly described |
|  |  |  | Primary paper | Other |  |  |
| 1 | Detailed description of the type of exercise equipment | Home-exercise book containing theoretical information about the management of kinesiophobia. | Pg. 233. |  | 1 |  |
| 2 | Detailed description of the qualifications, expertise and/or training | The patients underwent home-based treatment after being discharged from the Operative Unit of Physical Medicine and Rehabilitation of the Salvatore | Pg. 232-233. |  | 0 | Not clearly described |
|  |  | Maugeri Foundation's Scientific Institute in Lissone (Milan, Italy), a highly specialized rehabilitation centre in which physiatrists, physiotherapists, and psychologists highly experienced in knee pain and cognitive behavioural therapy treat more than 200 TKA patients every year. |  |  |  |  |
| 3 | Describe whether exercises are performed individually or in a group | Individually: Before returning home, the patients belonging to the experimental group were asked to continue the functional exercises learned during hospitalization... | Pg. 233. |  | 1 |  |
| 4 | Describe whether exercise are supervised or unsupervised; how they are delivered | Unclear during hospitalization and unsupervised after discharge (see above): They were also given a home-exercise book containing theoretical information about the management of kinesiophobia. | Pg. 233. |  | 0 | Not clearly described |
| 5 | Detailed description of how adherence to exercise is measured and reported |  |  |  | 0 | Not reported |
| 6 | Detailed description of motivation strategies | During the 6 months of home treatment, the patients received monthly phone calls from the rehabilitation team designed to strengthen treatment adherence. During the calls, the patients were also given an opportunity to ask for explanations about the concepts involved and, step by step, performed the required behavioural tasks.<br>In order to guarantee adhesion to the exercise and cognitive-behavioural principles and minimize the dropout rate, family doctors and relatives (spouses, significant others, or parent) were asked to support patient compliance actively during the course of the trial and inform staff promptly if any difficulty was encountered. | Pg. 233.<br>Pg. 233. |  | 1 |  |
| **7a** | Detailed description of the decision rule(s) for determining exercise progression | By a process of systematic self-desensitization, and once the patient could imagine and perform the scene without feeling fearful, the patient were asked to move on the next feared situation in the book. | Pg. 233. |  | 0 | Not clearly described. |
| **7b** | Detailed description of how the exercise program was progressed | See item 7a |  |  | 0 | Not clearly described |
| **8** | Detailed description of each exercise to enable replication | During their 15-day period of hospitalization, all of the enrolled patients underwent the same functional exercise-based rehabilitation program with the aim of improving knee range of motion, increasing knee and hip muscle strength, preventing thrombosis, and acquiring the most important functional strategies for ADL. The functional exercises included early walking training and how to use walking aids; functional task-oriented exercises, such as moving from a sitting to a standing position, ascending/descending stairs, and climbing obstacles (initially using a double handrail or parallel bars); weight-bearing exercises, such as walking in place and bilateral and unilateral knee flexion when standing; and other exercises aimed at recovering abilities, such as turning, sudden starts and stops, standing on unstable surfaces, and walking while changing speed and direction. Sessions of stationary cycling were added to optimize the strength and mobility of the knee. | Pg. 233. |  | 0 | Not clearly described |
| **9** | Detailed description of any home programme component | Before returning home, the patients belonging to the experimental group were asked to continue the functional exercises learned during hospitalization in twice-weekly 60-minute sessions for 6 months. | Pg. 233. |  | 0 | Not clearly described |
| **10** | Describe whether there are any non-exercise components | Experimental group: ... They were also given a home-exercise book containing theoretical information about the management of kinesiophobia, which was developed by our multidisciplinary rehabilitation team of physiatrists, clinical psychologists, and physiotherapists on the basis on the main situations avoided by the patients because of persistent knee pain, and the fear-avoidance beliefs emerging from discussion of their usual activities. | Pg. 233. |  | 1 |  |
| **11** | Describe the type and number of adverse events that occur during exercise | No information was collected concerning adverse effects. | Pg. 233. |  | 0 | Not reported |
| **12** | Describe the setting in which the exercises are performed | During the 6 months of *home treatment* | Pg. 233. |  | 1 |  |
| **13** | Detailed description of the exercise intervention | See item 8 |  |  | 0 | Not clearly described. |
| **14a** | Describe whether the exercises are generic (one size fits all) or tailored | See item 8 |  |  | 0 | Not clearly described. |
| **14b** | Detailed description of how exercises are tailored to the individual | See item 8 |  |  | 0 | Not clearly described. |
| 15 | Describe the decision rule for determining the starting level | By a process of systematic self-desensitization, and once the patient could imagine and perform the scene without feeling fearful, the patient were asked to move on the next feared situation in the book. | Pg. 233. |  | 1 |  |
| 16a | Describe how adherence or fidelity is assessed/measured |  |  |  | 0 | Not reported |
| 16b | Describe the extent to which the intervention was delivered as planned |  |  |  | 0 | Not reported |
| Total score |  |  |  |  | 6/19 |  |
| With inspiration from Slade et al. (2016): Consensus on Exercise Reporting Template (CERT): Explanation and Elaboration Statement. *Br J Sports Med* 2016;**50**:1428-1437. doi:10.1136/bjsports-2016-096651. |  |  |  |  |  |  |

